# Comorbid neuropathological diagnoses in early vs late-onset Alzheimer’s disease

**DOI:** 10.1101/2020.10.14.20213017

**Authors:** Salvatore Spina, Renaud La Joie, Cathrine Petersen, Amber L. Nolan, Deion Cuevas, Celica Cosme, Mackenzie Hepker, Ji-Hye Hwang, Zachary A. Miller, Eric J. Huang, Anna M. Karydas, Harli Grant, Adam L. Boxer, Maria Luisa Gorno-Tempini, Howard J. Rosen, Joel H. Kramer, Bruce L. Miller, William W. Seeley, Gil D. Rabinovici, Lea T. Grinberg

**Affiliations:** Memory and Aging Center, Department of Neurology, Weill Institute for Neurosciences, University of California, San Francisco, San Francisco, CA; Department of Pathology; University of California, San Francisco, San Francisco, CA; Department of Radiology and Biomedical Imaging; University of California, San Francisco, San Francisco, CA

## Abstract

Copathologies play an important role in the expression of the AD clinical phenotype and may influence treatment efficacy. Early-onset AD (EOAD), defined as manifesting before age 65, has been viewed as a relatively pure form of AD with a more homogenous neuropathological substrate. We sought to compare the frequency of common neuropathological diagnoses in a consecutive autopsy series of 96 patients with EOAD (median age of onset = 55 years, 44 females) and 48 with late-onset AD (LOAD) (median age of onset = 73 years, 14 females). The UCSF Neurodegenerative Disease Brain Bank database was reviewed to identify patients with a primary pathological diagnosis of AD. Prevalence and stage of Lewy body disease (LBD), limbic age-related TDP-43 encephalopathy (LATE), argyrophilic grain disease (AGD), hippocampal sclerosis (HS), cerebral amyloid angiopathy (CAA), vascular brain injury (VBI) and aging-related tau astrogliopathy (ARTAG) were compared between the two cohorts. We found at least one non-AD pathological diagnosis in 98% of patients with EOAD (versus 100% of LOAD), and the number of comorbid diagnoses per patient was lower in EOAD than in LOAD (median=2 versus 3, Mann-Whitney Z=3.00, p=0.002). LBD and CAA were common in both EOAD and LOAD (CAA: 86% versus 79%, Fisher exact p=0.33; LBD: 49% versus 42%, p=0.48, respectively), although amygdala-predominant LBD was more commonly found in EOAD than LOAD (22% versus 6%, p=0.02). In contrast, LATE (35% versus 8%, p<0.001), HS (15% versus 3%, p=0.02), AGD (58% versus 41%, p=0.052), and VBI (65% versus 39%, p=0.004) were more common in LOAD than EOAD, respectively. The number of copathologies predicted worse cognitive performance at the time of death on MMSE (1.4 points/pathology (95%CI [−2.5, −0.2]) and Clinical Dementia Rating – Sum of Boxes (1.15 point/pathology, 95%CI [0.45, 1.84]), across the EOAD and the LOAD cohorts. Prevalence of at least one ApoE e4 allele was similar across the two cohorts (52%) and was associated with a greater number of copathologies (+0.42, 95%CI [0.01, 0.82], p=0.04), independent of age of symptom onset. Our findings suggest that non-AD pathological diagnoses play an important role in the clinical phenotype of EOAD with potentially significant implications for clinical practice and clinical trials design.

## Introduction

The coexistence of multiple proteinopathies and vascular brain injury (VBI) is a common finding in the brains of elders, the number of distinct pathological entities being strongly correlated with age and genetic factors, such as the ApoE genotype (Karanth et al., 2020; Robinson et al., 2018; Suemoto et al., 2017). While numerous proposed mechanisms of mutual induction support the biological interaction of some of these proteinopathies, the occurrence of other copathologies appears to be a time-dependent phenomenon, with the likelihood of accumulating more distinct types of brain pathology reflecting longer survival (Coulthard and Love, 2018). Alzheimer’s disease (AD) neuropathological changes (ADNC) are frequently associated with a high prevalence of coexistent Lewy body disease (LBD) and/or TDP-43 proteinopathy, which influence the clinical phenotype (Higashi et al., 2007; James et al., 2016; McAleese et al., 2017; Nelson et al., 2019; Swirski et al., 2014). While prior studies have begun elucidating the prevalence of coexisting pathology in AD, differences in the number and type of copathologies between sporadic early-onset AD (EOAD) and late-onset AD (LOAD) patients are less well characterized (Robinson et al., 2018; Suemoto et al., 2017). Sporadic early-onset AD (EOAD) defines the small proportion of patients with AD whose clinical onset occurs before the age of 65 years, in the absence of AD pathogenic mutations. Despite the historical and arbitrary age-based classification (Fox and Phil, 1986), and in spite of sharing the same hallmark neuropathological features, beta-amyloid plaques and tau-immunoreactive neurofibrillary tangles (NFT), EOAD and LOAD are commonly referred to as potentially diverging clinicopathological entities on the basis of differences in the phenotypic presentation and genetic predisposition (Mendez, 2019). Because of the onset in young age, EOAD represents the ideal model to contrast the two hypotheses of the mutual induction of distinct copathologies vs. the age-dependent model leading to copathology accumulation in AD. Here, we report the frequency and severity of coexisting copathologies in a cohort of consecutive patients with a primary neuropathological diagnosis of AD, and at the age of onset of cognitive symptoms consistent with either EOAD or LOAD.

## Methods

### Neuropathological criteria for cases selection

We searched the Neurodegenerative Diseases Brain Bank (NDBB) database of the University of California, San Francisco to identify cases with a primary pathological diagnosis of AD, assessed by expert neuropathologists (SS, EJH, WWS, and LTG) between September 2008 and March 2020. The brains were obtained postmortem, processed, and analyzed according to the NDBB research protocol, as previously described (Spina et al., 2019). Out of 516 cases, the search returned 190 cases with primary pathological diagnosis of Alzheimer’s disease. Primary pathological diagnosis is herein defined as the most developed neuropathological entity, which severity and regional distribution are thought to explain the majority of the patient’s clinical cognitive and behavioral phenotype. Contributing pathological diagnosis is defined as a coexistent neuropathological entity that is sufficiently developed to likely contribute to the primary clinical phenotype or to additional clinical features that cannot be otherwise attributed to the primary pathological diagnosis. Since the goal of the study was to ascertain the relative frequency of the most common non-AD neurodegenerative copathologies (LBD, LATE, argyrophilic grain disease (AGD), hippocampal sclerosis (HS), and cerebral amyloid angiopathy (CAA), among the two cohorts of EOAD and LOAD, 12 cases were excluded because of a co-primary or contributing diagnosis of frontotemporal lobar degeneration (FTLD) (Dickson et al., 1994; Ferrer et al., 2008; Mackenzie et al., 2010; McKeith et al., 2017; Nelson et al., 2019). CAA was added to the total number of non-AD copathologies because its presence is not accounted in the current pathological diagnostic criteria for ADNC, and because of a possible direct impact on cognition independent of CAA-related vascular brain injury (VBI) or CAA-related neuroinflammation (CAA-RI) (Banerjee et al., 2017; Montine et al., 2012). Five cases were excluded because of a co-primary diagnosis of primary lateral sclerosis with TDP-43 inclusions, CAA-RI, chronic traumatic encephalopathy (CTE), or VBI, and one additional case was excluded because of contributing traumatic brain injury with diffuse axonal degeneration (Dickson et al., 2007; Kirshner and Bradshaw, 2015; McKee et al., 2013). CAA-RI was excluded because of the subacute neurobehavioral symptoms, partially responsive to immunosuppression, that differentiates this syndrome from the clinical course of the other AD co-pathologies (Kirshner and Bradshaw, 2015). This led to the identification of a pathological cohort of 173 cases.

Next, cases were excluded if information pertinent to Alzheimer’s Disease Neuropathological Changes (ADNC) severity, including Thal phase, Braak Stage, and CERAD neuritic plaque frequency stage were unavailable (Montine et al., 2012). This led to the exclusion of 20 additional cases, and the remaining of 153 cases. Then, we excluded cases lacking cognitive decline and cases with an autosomal dominant AD (see below), resulting in a final cohort of 144. Fig. 1 depicts the flowchart of case selection.

**Figure 1.**
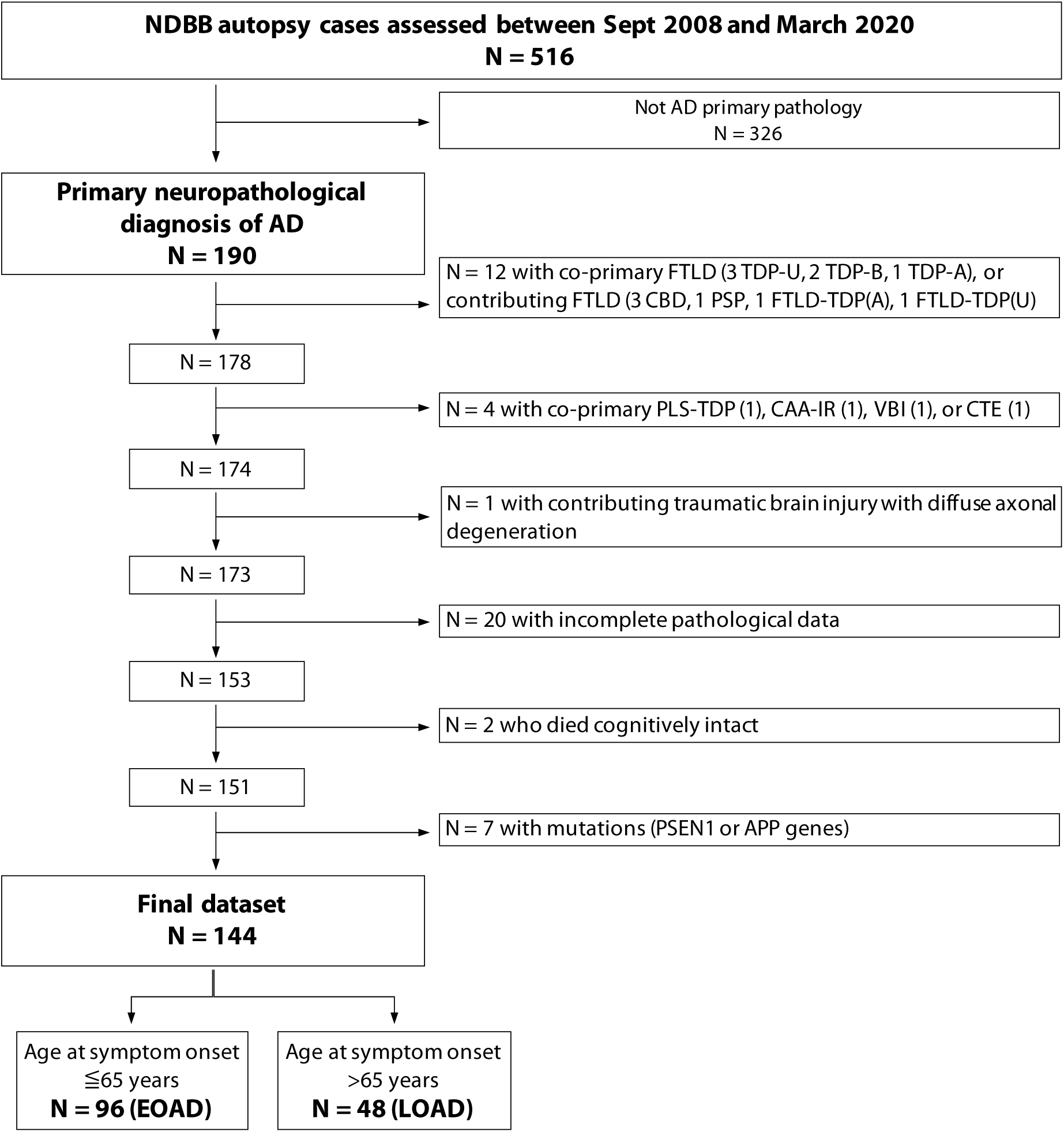
Flowchart of patient selection.

### Additional neuropathological assessment

LBD staging (Braak 1-6) was assessed through alpha-synuclein immunohistochemistry according to published criteria (Braak et al., 2003). Because of the high prevalence of cases with LBD pathology non-conforming to the Braak criteria and primarily confined to the amygdala and/or limbic brain regions with minimal or no involvement of brainstem and neocortex, an additional category of amygdala-predominant LBD was considered. TDP-43 proteinopathy was assessed as absent, limbic-predominant (LATE-NC stages I and II) or diffuse (LATE-NC stage III) according to published criteria (Nelson et al., 2019). Argyrophilic grain disease (AGD) was assessed as previously described (Rodriguez et al., 2016), and staged as absent, limbic-predominant, or diffuse/neocortical. A diagnosis of HS was given to cases with above 90% neuronal loss in CA1/subiculum (Nelson et al., 2013). VBI pathology was considered present when either macroinfarcts (either territorial or lacunar) or old microinfarcts (areas of regional ischemia with tissue loss only visible through microscopic assessment) were observed. CAA was assessed as absent, mild (limited to the leptomeningeal vessels), moderate (sparsely involving intraparenchymal vessels), or severe (involving the majority of intraparenchymal arterioles with circumferential pattern of deposition within the vessel walls) (Grinberg and Thal, 2010).

Aging-related tau astrogliopathy (ARTAG) (Kovacs et al., 2016), including argyrophilic thorny astrocytes in clusters (ATAC) pathology, also known as white-matter thorny shaped astrocytes (Munoz et al., 2007; Nolan et al., 2019), was assessed. Still, its presence was not added to the total number of copathologies since data were missing for 36 participants. Presence of contributing chronic traumatic encephalopathy (CTE) was also assessed (McKee et al., 2013). Since CTE is linked to exposure to repeated head trauma within a specific environmental setting, this pathology was not added to the total number of copathologies to avoid ascertainment bias between the two cohorts. Except for data on ATAC (missing in one case) and ARTAG (missing on 35 cases), only cases (n=153, including 103 with EOAD and 50 with LOAD) for which the full assessment of the aforementioned pathological entities was carried out were included in the study. Taking advantage of data obtained through a previous study (Petersen et al., 2019), NFT densities were available from the middle frontal gyrus, superior temporal gyrus, primary motor cortex, angular gyrus, and/or CA1/subiculum of 54 EAOD and 23 LOAD patients.

### Clinical assessment, genetic screening, and cohort definitions

Clinical information on each of the 153 patients in the pathology dataset was reviewed. All cases had been enrolled during life in longitudinal studies on neurodegenerative dementia at the Memory and Aging Center, UCSF. Information on sex, age at onset, and age at death were collected. Clinical diagnosis were provided by expert behavioral neurologists, according to published criteria (Armstrong et al., 2013; Crutch et al., 2017; Geschwind, 2016; Gorno-Tempini et al., 2011; McKeith et al., 2017; McKhann et al., 2011; Montenigro et al., 2014; Postuma et al., 2015; Rascovsky et al., 2011). For this study’s purpose, the reported clinical diagnosis for each patient refers to the final best-fit clinical syndromic diagnosis at the latest UCSF clinical assessment. Age at onset was defined as the age of occurrence of the first clinical symptom of AD as determined by the UCSF clinician based on the patient’s clinical history. Patients with age at onset of 65 years or younger were assigned to the EOAD group, while patients with age at onset higher than 65 were assigned to the LOAD group. Age at onset was not available for three participants: two of them died cognitively unimpaired respectively at age 91 and 97 and were excluded from the study; the third subject developed mild cognitive impairment a few years before dying at age 74 and was therefore included in the LOAD group. Genetic analyses were carried out in 137/153 patients as previously described (Perry et al., 2017). Genetic screening for pathogenic mutations in the *APP, PSEN1, PSEN2, C9ORF72, GRN, MAPT, FUS*, and *TARDBP* genes were carried out in 133 patients. Two additional patients were only screened for the *C9ORF72* pathogenic expansion and two more patients were only screened for mutations in either *C9ORF72, MAPT* or *GRN*. These studies led to the identification of 5 carriers of an *APP* mutation and 2 carriers of a *PSEN1* mutation. These 7 patients were excluded from the study. This led to a final dataset of 144 patients, subdivided into an EOAD cohort of 96 patients and a LOAD cohort of 48 participants, whose data were used for the analyses. Clinical progression was assessed using consecutive scores on Mini Mental State Exam (MMSE) available for 137 patients (382 observations), and Clinical Dementia Rating-Sum of Boxes (CDR-SoB) available for 140 patients (485 observations). ApoE genotype was available for 91 EOAD and 42 LOAD subjects).

### Statistical analyses

Frequency of NFT Braak stages, beta-amyloid Thal phases, CAA stages, LBD Braak stages and presence or absence of amygdala-predominant LBD, TDP-43 proteinopathy severity, HS, AGD, ATAC, and ARTAG pathology were compared between the two groups of EOAD and LOAD. The number of co-pathologies is defined by the presence of LBD, TDP-43 proteinopathy, HS, AGD, CAA and/or VBI. Comparison of ordinal variables (e.g., NFT Braak stage) was carried out using Mann-Whitney tests. Comparison of binarized (presence or absence) measures for any pathological entity between the two groups was assessed using the Fisher exact test. Chi-square test was used to assess differences in the frequency of LBD subtypes (i.e., absent; conforming to PD Braak stage; or amygdala-predominant only) between groups. For some complementary analyses, age of onset was treated as a continuous variable and we assessed its association with neuropathological measures using Spearman’s rho coefficient (for continuous or ordinal measures of neuropathology), Mann-Whitney U tests (for binary pathology measures), or Kruskal-Wallis tests (for non-ordinal neuropathology categories with more than 2 levels). For Mann-Whitney tests, we report the results using the common language effect size measure (U / (n_1_*n_2_), representing the probability that a random value from Group1 is greater than a random value from Group2, similar to the area under the receiver operating curve.

Logistic regression analyses were used to assess how age and ApoE e4 carrier status predicted the presence of each co-pathology while a multiple regression model was used to assess the association between age and ApoE e4 on the number of copathologies. Significance values are reported with a p < 0.05.

To assess the associations between age of disease onset, number of copathologies, and the rate of cognitive and clinical decline, we tested a series of linear mixed effect models using either MMSE or CDR-SoB as the dependent variable. All models included a fixed effect for time until death (in years) and random intercept and slope; the time variable was not centered so that the model intercept indicates the predicted MMSE or CDR-SoB value at time of death. Additional fixed effects were considered including main effects for cohort (EOAD versus LOAD) and number of copathologies (0 to 6), and three interactions: time until death * cohort, time until death * number of copathologies, and a triple time until death * cohort * number of copathologies interaction. For each of the two clinical measures, all 14 potential models (see Supplementary Tables 2 and 3) were considered, and the optimal model was chosen based on the minimal Akaike Information Criterion (AIC) and Bayesian Information Criterion (BIC).

## Data availability

All data used in this study are available for review upon request.

## Results

### Demographics and clinical presentation

Age at onset was 55.2 ± 5.8 (mean ± SD), median = 55, min = 44, max = 65 years in the EOAD group (n=96) and 74.0 ± 5.7 years, median = 73, min = 66, max = 90 in the LOAD group (n=48). Age at death was 66.4 ± 6.6 years in the EOAD group, and 83.0 ± 5.9 years in the LOAD group. Disease duration was longer in the EOAD group (11.2 ± 3.9 years) than in the LOAD group (9.0 ± 3.4 years, t = 3.3, p = 0.001, Cohen’s d = 0.59). Both groups included a majority of men (54% in EOAD, 71% in LOAD, Fisher exact p = 0.07). In the EOAD group, 57 (59%) patients were diagnosed with amnestic AD, 11 with posterior cortical atrophy (PCA), 10 with logopenic variant primary progressive aphasia (lvPPA), 7 with corticobasal syndrome (CBS), 3 with behavioral variant frontotemporal dementia (bvFTD), 2 with mild cognitive impairment, 2 with dementia with Lewy bodies (DLB), 1 with Parkinson’s disease dementia, 1 with nonfluent variant PPA (nfvPPA), 1 with unspecified PPA and 1 with unspecified rapidly progressive dementia. In the LOAD group 33 (69%) patients were diagnosed with amnestic AD, 5 with MCI, 3 with lvPPA, 2 with CBS, 1 with PCA, 1 with bvFTD, 1 with LBD, 1 with traumatic encephalopathy syndrome, and 1 patient did not meet research diagnostic criteria.

### Severity of Alzheimer’s disease pathological features is higher in EOAD than LOAD

NFT Braak stage (Mann-Whitney U = 2860, Z = 4.32, p < 0.001) and beta-amyloid Thal phase (U = 2724.5, Z = 3.10, p = 0.002) were significantly higher in the EOAD group compared to the LOAD group (Fig. 2A). When AD pathological stages were correlated with age of onset as a continuous variable within the whole cohort (Supplementary Figure 1A), we observed significant reduction of NFT Braak stage (rho = −0.3, p < 0.001) and Thal phase (rho = −0.30, p < 0.001) with age.

**Figure 2.**
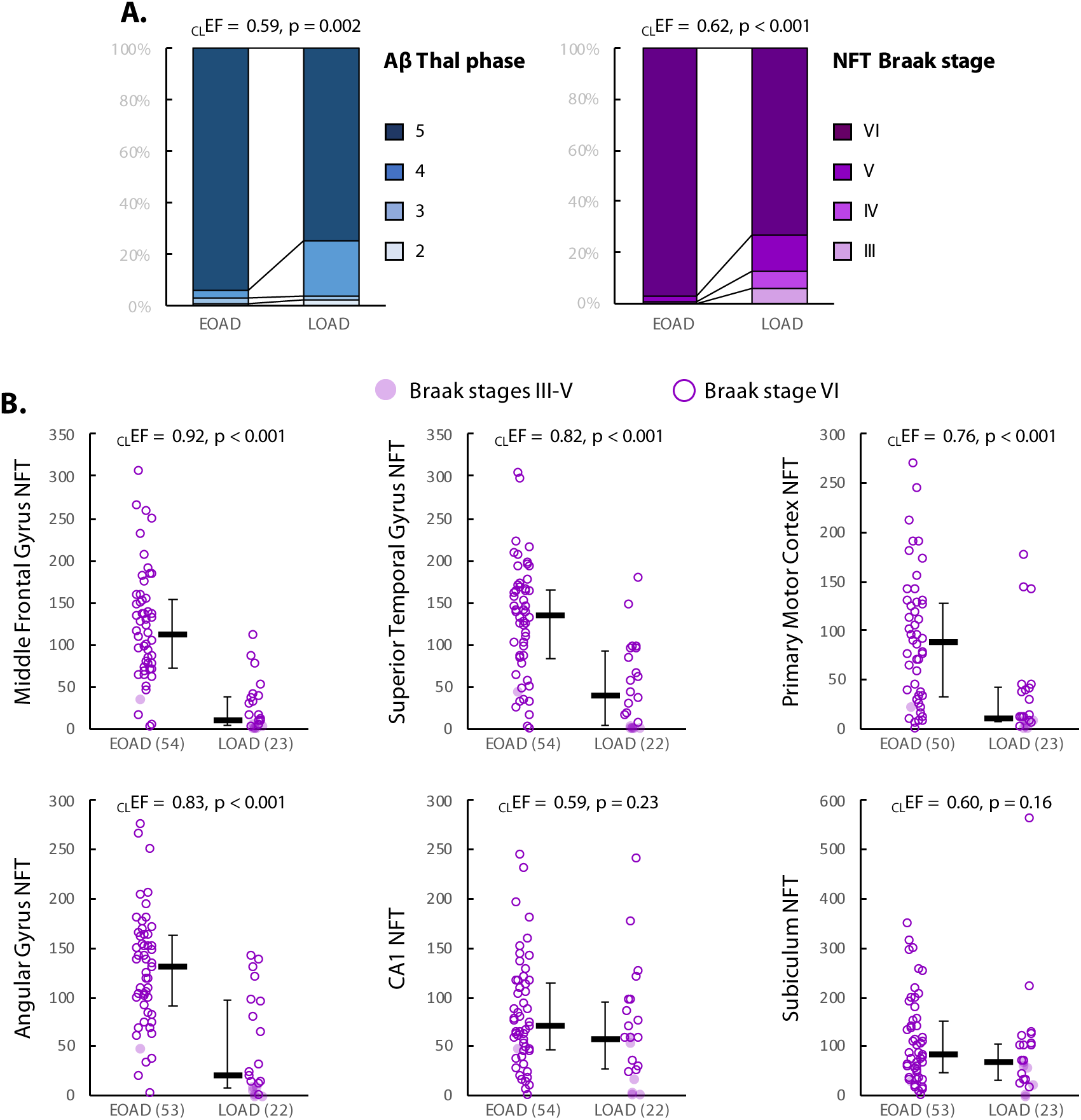
Alzheimer’s disease pathology in EOAD and LOAD. A. Ordinal neuropathological scales. B. quantitative analyses of NFT density in 6 predefined regions (density/mm3); bars indicate median and interquatile range. For all variables, Mann-Whitney U tests were conducted to compare the two groups; common language effect sizes (CLEF=U/(nEOAD*nLOAD)) represent the probability that a random value from the EOAD group is greater than a random value from the LOAD group. nEOAD = 96 and nLOAD = 48, unless otherwise specified.

Quantitative analyses of NFT density in a subgroup of the cohort showed strong group differences between EOAD and LOAD (Fig. 2B). Compared to LOAD, patients with EOAD had higher NFT density in all investigated cortical areas: middle frontal gyrus (U = 1143.5, Z = 5.81, p < 0.001), superior temporal gyrus (U = 968.5, Z = 4.28, p < 0.001), primary motor cortex (U = 878.5, Z = 3.60, p < 0.001), and angular gyrus (U = 971, Z = 4.51, p < 0.001). Differences were milder and did not reach the *α* < 0.05 significance threshold in the two hippocampal regions examined: CA1 (U = 699.5, Z = 1.20, p = 0.23) and subiculum (U= 703.5, Z = 1.40, p = 0.16). Results were unchanged when analyses were restricted to cases with Braak stage VI or when group differences were tested using analyses of covariance controlling for disease duration and sex (see Supplementary Table 1). Similar associations were observed when considering age of onset as a continuous variable (Supplementary Figure 1B).

### Coexistent non-AD pathologies are common in EOAD, and their prevalence increases with age along the AD spectrum

The number of non-AD coexistent pathologies was significantly higher in the LOAD group compared to the EOAD (median = 3 versus 2, U = 1619, Z = 3.00, p = 0.002; see Fig. 3A). This relationship was also observed when age of onset was treated as a continuous variable in the whole cohort (rho = −0.26, p < 0.001; Fig. 3B). Coexistent non-AD pathologies were found in all patients with LOAD (48/48) and in 98% (94/96) of EOAD cases; 69 of these 96 cases had more than one non-AD copathology. Among the 25 EOAD cases with single coexisting pathology, 19 showed coexistent CAA pathology, 3 coexistent AGD, and 3 concurrent vascular brain injury. Two non-AD copathologies were observed in 31/96 EOAD patients, three coexistent pathological entities in 26/96 EOAD patients, four in 9/96 patients, five in 2/9<u>6</u> patients, and six non-AD coexistent pathologies in 1/96 EOAD patients. The overall prevalence of coexistent pathologies in the EOAD group was as follows: CAA 86%, LBD 49%, AGD 41%, vascular brain injuries 39%, TDP-43 proteinopathy 8%, and hippocampal sclerosis 3% (Fig. 4). In the LOAD group, the prevalence of non-AD copathology was as follows: CAA 79%, VBI 65%, AGD 58%, LBD 42%, TDP-43 35%, and HS 15%. Two subjects in the LOAD group (4%) had concurrent CTE pathology. They both had a history of repetitive head trauma secondary to professional participation in a contact sport.

**Figure 3.**
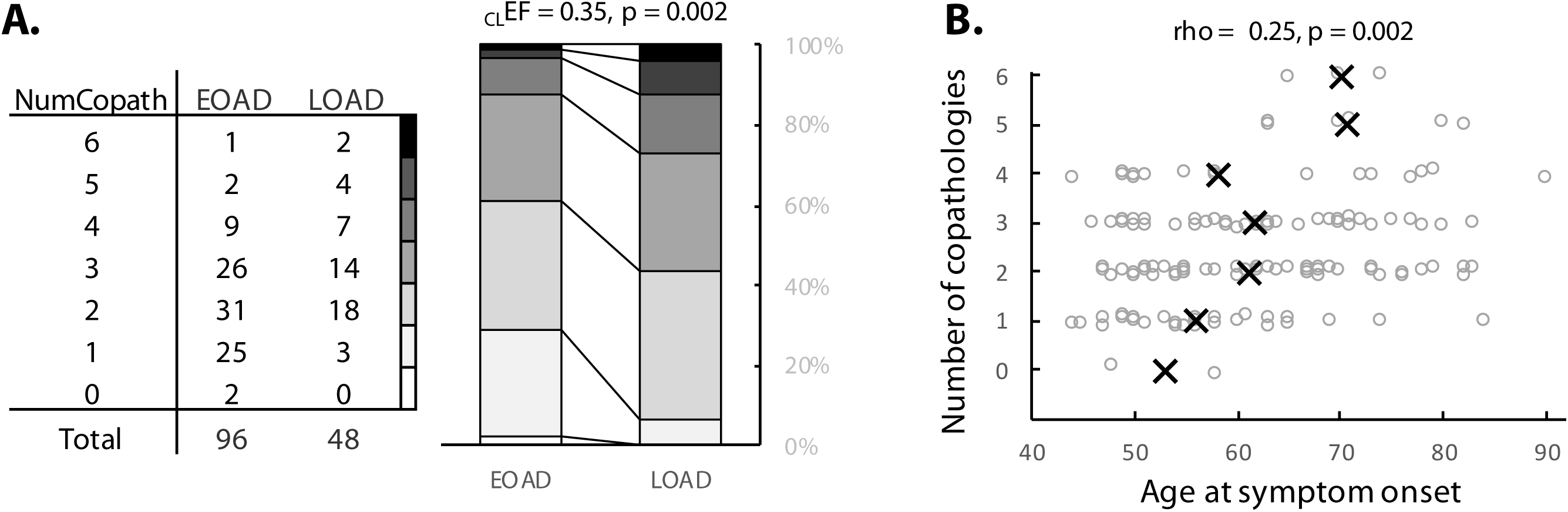
Number of coexistent pathologies. A. The table indicates the numbers of EOAD/LOAD patients with respective total number of copathologies, while the stacked bars illustrate the higher number of copathologies in the LOAD group. Mann-Whitney U test were conducted to compare the two groups; common language effect size (CLEF=U/(nEOAD*nLOAD)) represents the probability that a random value from the EOAD group is greater than a random value from the LOAD group. A Fisher exact test was also run to compare the proportion of copathology-free cases between the two groups. B. Analyses conducted with age of onset as a continuous variable. Black crosses indicate the median ages for each level of copathology; a random jitter was applied on the y axis to visualize all individual datapoints

**Figure 4.**
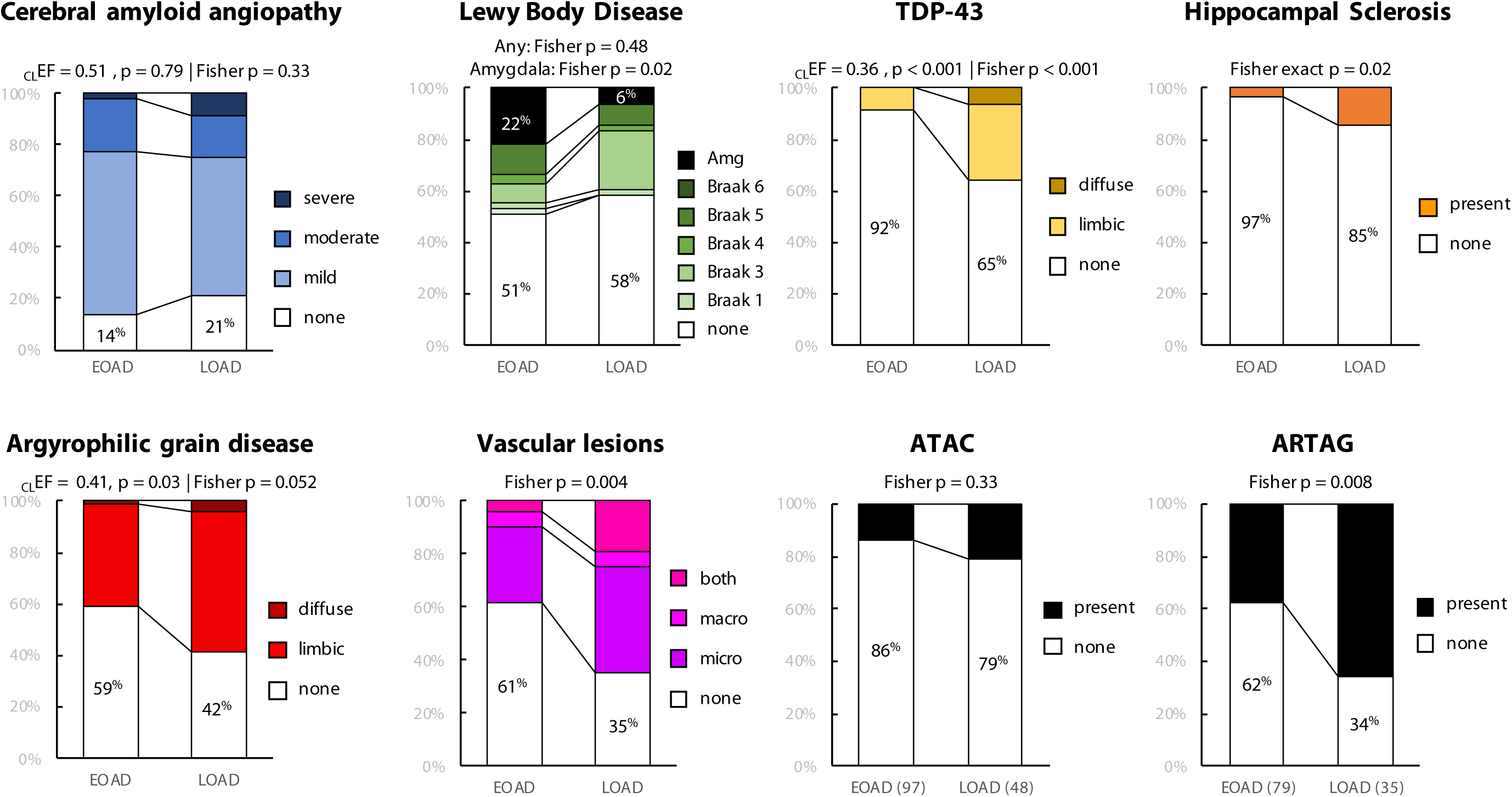
Details of coexistent non-AD pathologies. For all copathologies, Fisher exact tests were run to compare the proportion of copathology-free cases between the two groups. For Lewy body disease, an additional Fisher exact test was run to compare the frequency of amygdala-predominant patterns between the two groups. For ordinal variables (TDP-43, AGD), Mann-Whitney U tests were conducted to compare the two groups; common language effect sizes (CLEF=U/(nEOAD*nLOAD)) represent the probability that a random value from the EOAD group is greater than a random value from the LOAD group. nEOAD = 96 and nLOAD = 48, unless otherwise specified

### CAA pathology is very common in both EOAD and LOAD

CAA frequency was similar in the EOAD and LOAD groups (86% versus 79%, Fisher exact p = 0.33, Fig. 4); no group difference was observed when considering the severity of CAA as an ordinal variable (U = 2360, Z = 0.27, p = 0.79), or when considering age of onset as a continuous variable (Supplementary Figure 2).

### Lewy-body pathology is equally prevalent in EOAD and LOAD, and more commonly amygdala-predominant type in EOAD

We found no difference in the presence of alpha-synuclein copathology among the two groups: 47/96(49%) in EOAD versus 20/48 (42%) in LOAD (Fisher exact p = 0.48, Fig. 4). However, when LBD was assessed as three distinct subtypes (i.e. absent; conforming to PD Braak stage; or amygdala-predominant only), we observed a difference between the EOAD and LOAD groups (χ^2^ = 5.75, p=0.056) primarily driven by the higher proportion of amygdala-predominant LBD in EOAD (21/96, 22%) compared to LOAD (3/48, 6%); Fisher exact p = 0.02. Results were consistent when analyzing age of onset as a continuous variable (Supplementary Figure 2).

### TDP-43 proteinopathy, AGD, hippocampal sclerosis, aging-related tau astrogliopathy (ARTAG) and vascular brain injuries were more frequent and/or severe in LOAD compared to EOAD

Coexistent TDP-43 proteinopathy (Fisher exact p < 0.001), hippocampal sclerosis (Fisher exact p = 0.02) and AGD (Fisher exact p = 0.052) were more prevalent in the LOAD group compared to EOAD (see Fig. 4). The prevalence of coexistent vascular lesions was higher in LOAD than EOAD (65% versus 39%, Fisher exact p = 0.004); post hoc analyses showed that both microinfarct (Fisher exact p = 0.004) and macroscopic (Fisher exact p = 0.02) lesions were more frequent in LOAD than EOAD.

ATAC pathology was more commonly seen in LOAD (21%) than EOAD (14%), though the difference did not reach statistical significance (Fisher exact p = 0.33). ARTAG pathology, not inclusive of ATAC, was significantly more common in LOAD (66%) than EOAD (38%), (Fisher exact p = 0.008).

Similar results were observed when considering age as a continuous variable (Supplementary Figure 2).

### Influence of ApoE e4

An ApoE e4 allele was found in 47/91 (52%) EOAD subjects and in 22 (52%) LOAD patients. The effect of APOE4 (coded as ApoE e4 carriers versus non-carrier) on the presence of coexisting pathologies was tested through regression models, always including age of onset as a covariate based on previous results. Additional models were run with sex and disease duration as additional predictors but resulted in comparable conclusions. A multiple linear regression model predicting the number of copathologies showed that older age of onset (estimate = 0.0350, 95%CI [0.0154, 0.0545], standardized estimate = 0.293, p = 0.0006) and APOE4 (estimate = 0.4163, 95%CI [0.0117, 0.8208], standardized estimate = 0.336, p = 0.044) were independently associated with a higher number of copathologies. A series of exploratory binomial logistic regression models were run to determine whether APOE4 status was associated with the presence of a specific copathology (all coded as present versus absence regardless of severity/stage); age of onset was included in all models. Detailed results are available in Tab. 1. No significant association was found between APOE4 status and CAA, alpha-synuclein, TDP-43, hippocampal sclerosis, vascular lesions, ATAC, or ARTAG (p’s *≥* 0.10). A trend toward statistical significance was observed for the association with AGD (p = 0.08). APOE4 status was not associated with beta-amyloid Thal phase (U = 2228.5, Z = 0.11, p = 0.91), NFT Braak stage (Mann-Whitney U = 2156.5, Z = 0.77, p = 0.44), or regional NFT density (Supplementary Table 2).

**Table 1.**
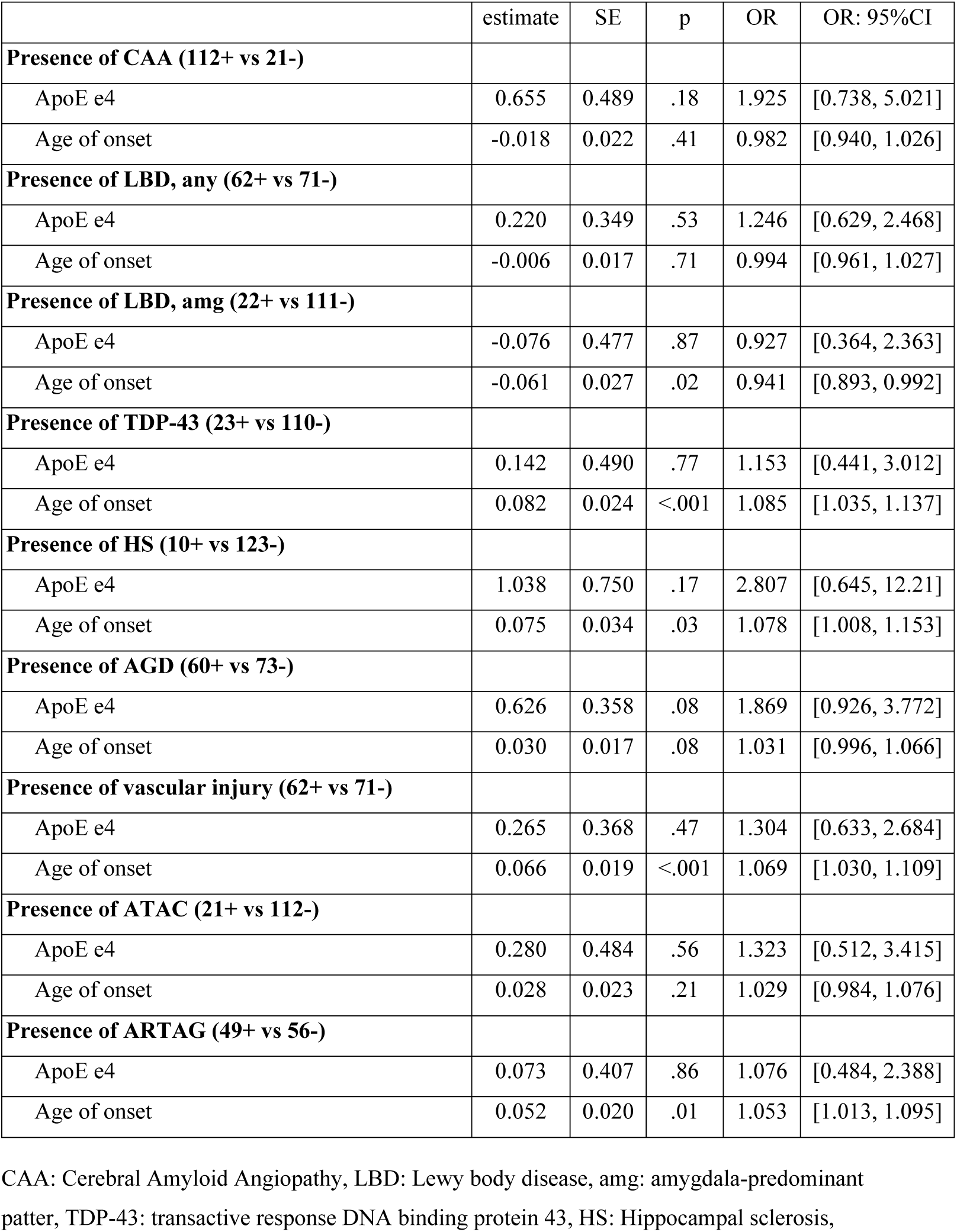
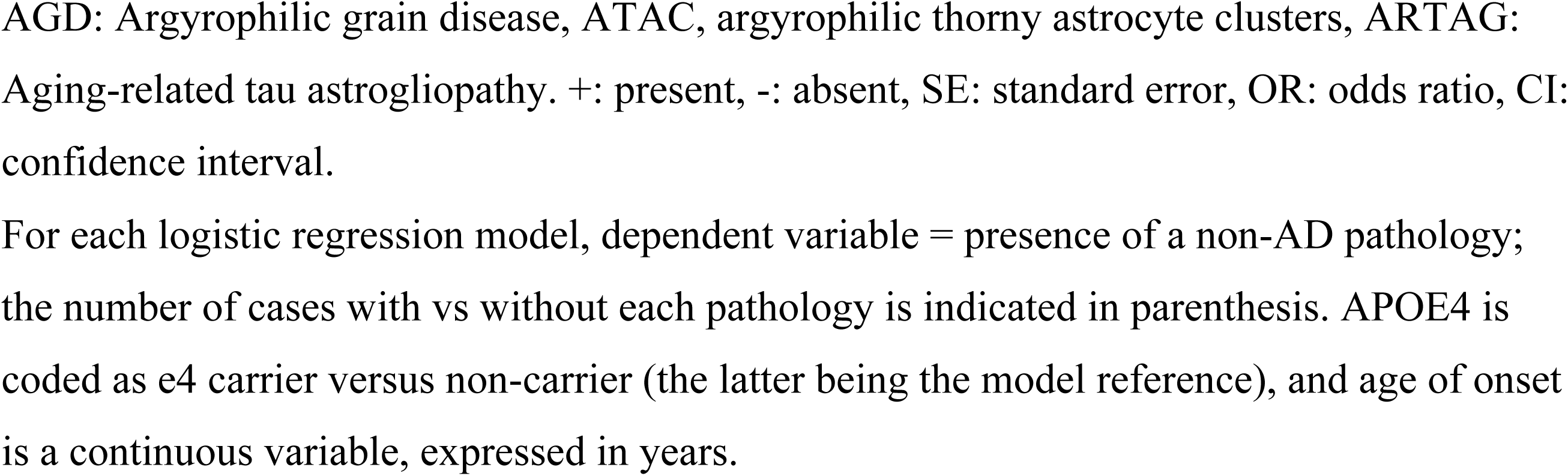
Logistic regressions testing the relative impact of ApoE e4 and age of onset on the presence of each copathology.

|  | estimate | SE | p | OR | OR: 95%CI |
| --- | --- | --- | --- | --- | --- |
| **Presence of CAA (112+ vs 21-)** |  |  |  |  |  |
| ApoE e4 | 0.655 | 0.489 | .18 | 1.925 | [0.738, 5.021] |
| Age of onset | -0.018 | 0.022 | .41 | 0.982 | [0.940, 1.026] |
| **Presence of LBD, any (62+ vs 71-)** |  |  |  |  |  |
| ApoE e4 | 0.220 | 0.349 | .53 | 1.246 | [0.629, 2.468] |
| Age of onset | -0.006 | 0.017 | .71 | 0.994 | [0.961, 1.027] |
| **Presence of LBD, amg (22+ vs 111-)** |  |  |  |  |  |
| ApoE e4 | -0.076 | 0.477 | .87 | 0.927 | [0.364, 2.363] |
| Age of onset | -0.061 | 0.027 | .02 | 0.941 | [0.893, 0.992] |
| **Presence of TDP-43 (23+ vs 110-)** |  |  |  |  |  |
| ApoE e4 | 0.142 | 0.490 | .77 | 1.153 | [0.441, 3.012] |
| Age of onset | 0.082 | 0.024 | <.001 | 1.085 | [1.035, 1.137] |
| **Presence of HS (10+ vs 123-)** |  |  |  |  |  |
| ApoE e4 | 1.038 | 0.750 | .17 | 2.807 | [0.645, 12.21] |
| Age of onset | 0.075 | 0.034 | .03 | 1.078 | [1.008, 1.153] |
| **Presence of AGD (60+ vs 73-)** |  |  |  |  |  |
| ApoE e4 | 0.626 | 0.358 | .08 | 1.869 | [0.926, 3.772] |
| Age of onset | 0.030 | 0.017 | .08 | 1.031 | [0.996, 1.066] |
| **Presence of vascular injury (62+ vs 71-)** |  |  |  |  |  |
| ApoE e4 | 0.265 | 0.368 | .47 | 1.304 | [0.633, 2.684] |
| Age of onset | 0.066 | 0.019 | <.001 | 1.069 | [1.030, 1.109] |
| **Presence of ATAC (21+ vs 112-)** |  |  |  |  |  |
| ApoE e4 | 0.280 | 0.484 | .56 | 1.323 | [0.512, 3.415] |
| Age of onset | 0.028 | 0.023 | .21 | 1.029 | [0.984, 1.076] |
| **Presence of ARTAG (49+ vs 56-)** |  |  |  |  |  |
| ApoE e4 | 0.073 | 0.407 | .86 | 1.076 | [0.484, 2.388] |
| Age of onset | 0.052 | 0.020 | .01 | 1.053 | [1.013, 1.095] |

CAA: Cerebral Amyloid Angiopathy, LBD: Lewy body disease, amg: amygdala-predominant pattern, TDP-43: transactive response DNA binding protein 43, HS: Hippocampal sclerosis,
AGD: Argyrophilic grain disease, ATAC, argyrophilic thorny astrocyte clusters, ARTAG: Aging-related tau astrogliopathy. +: present, -: absent, SE: standard error, OR: odds ratio, CI: confidence interval.
For each logistic regression model, dependent variable = presence of a non-AD pathology; the number of cases with vs without each pathology is indicated in parenthesis. APOE4 is coded as e4 carrier versus non-carrier (the latter being the model reference), and age of onset is a continuous variable, expressed in years.

### Analyses of cognitive and clinical decline

For both MMSE and CDR-SoB, the optimal linear fixed effect model included three main fixed effects (time until death, cohort group, and number of copathologies), and the interaction between time until death and cohort group (Supplementary Table 3 for MMSE, Supplementary Table 4 for CDR-SoB), which were all significant (see Fig. 5B and Supplementary Figure 3A for full description of each model).

**Figure 5.**
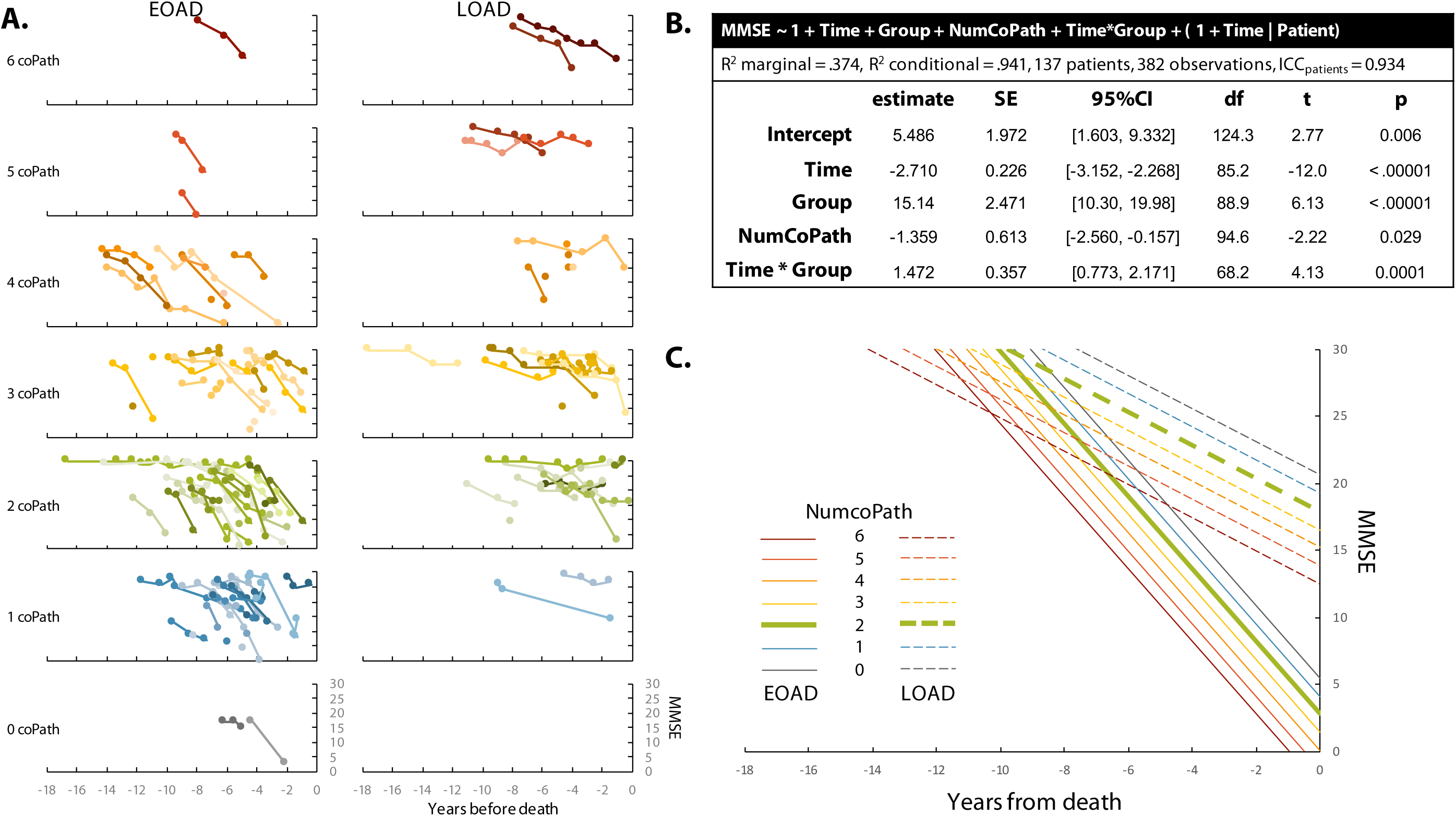
Ante mortem cognitive decline based on MMSE. A. Individual trajectories in MMSE scores (raw values), stratified by age of onset group (columns) and number of copathologies (rows). Color variations were randomly assigned to help distinguish overlapping lines.B. Optimal linear mixed effect model as identified in eTable3.C. MMSE trajectories as predicted by model in B. The trajectories for EOAD and LOAD patients with 2 copathologies (the median number of copathologies in the whole cohort) are emphasized by a thicker line.

Briefly, the MMSE model (137 patients, 382 observations, n=40 patients have only 1 timepoint, number of available timepoints per patients: median = 2, min-max = 1-12, years between first and last timepoints: median = 1.5, min-max = 0-12.2) showed a difference between the EOAD and LOAD cohorts, with the former being more severely impaired (mean difference = 15.1, 95%CI [10.3, 20.0]) at time of death. Each additional copathology was associated with a decrease of 1.4 MMSE point (95%CI [−2.5, −0.2]) at time of death. The time * age of onset cohort interaction was highly significant (p = 0.0001) and indicated a greater antemortem decline in MMSE in EOAD (−2.7 point / year, 95%CI [−3.2, −2.3]) compared to LOAD (−1.2, 95%CI [−1.8, −0.7]).

Results were similar with CDR-SoB (140 patients, 485 observations, (n=22 patients have only 1 timepoint, number of available timepoints per patients: median = 3, min-max = 1-10, years between first and last timepoints: median = 2.4, min-max = 0-11.5) (Supplementary Figure 3), with the EOAD group showing higher CDR-SoB scores than the LOAD at time of death (mean difference = 5.95, 95%CI [3.41, 8.48]), and each additional copathology being associated with greater CDR-SoB (1.15, 95%CI [0.45, 1.84]) at death. The time * group interaction (p = .04) also indicated a greater premortem rate of CDR-SoB increase in EOAD (1.47 point / year, 95%CI [1.25, 1.68]) compared to LOAD (1.09, 95%CI [0.80, 1.37]).

Although none of the linear mixed effect models selected based on the lowest AIC/BIC indices included an interaction term between time and number of copathologies, complementary analyses showed that when these terms were forced into the models (i.e. using model 10 from Supplementary Tables 3-4), they were not significant : p = 0.72 for the MMSE model and p = 0.09 for the CDR-SB model.

## Discussion

Frequency and severity of brain copathologies increase with age and contribute to the severity of the clinical phenotype in individuals with cognitive impairment (Karanth et al., 2020; Robinson et al., 2018; Suemoto et al., 2017). The co-occurrence of multiple distinct neuropathological features in younger patients with dementia has been considered a rare phenomenon. AD is the most common neuropathological substrate of dementia in both the older and younger populations. While the hallmark AD neuropathological end-stage features of EOAD and LOAD are similar, several aspects of clinical presentation and genetic susceptibility have suggested the existence of fundamental biological differences between EOAD and LOAD that go beyond the arbitrary and simplistic age-related categorization (Tellechea et al., 2018). The coexistence of distinct non-AD pathologies in individuals with EOAD has so far eluded proper ascertainment because of the limited availability of large autopsy cohorts of EOAD subjects. Our study shows the presence of one copathology in a quarter of patients with EOAD, two in about a third, three in more than a quarter, and four in 9% of our cohort. Our data also show an important cumulative effect of the number of coexistent non-AD pathologies on clinical progression rate, a feature of both EOAD and LOAD. These data have important implications for the enrollment of EOAD subjects in longitudinal studies and clinical trials, suggesting that non-AD pathological diagnoses are common and consequential, even in younger patients with AD.

Coexistent LBD pathology significantly worsens the severity of the clinical presentation of AD (Malek-Ahmadi et al., 2019). LBD pathology is frequent and clinically significant in autosomal dominant AD, typically manifesting with an early age of onset (Leverenz et al., 2006). Our study shows that LBD is as prevalent in sporadic EOAD as it is in LOAD, though in about a quarter of EOAD patients LBD copathology is predominantly confined to the amygdala. While the significance of amygdala-predominant LBD in determining the AD clinical phenotype is likely limited (Roudil et al., 2018; Sorrentino et al., 2019; Uchikado et al., 2006), recent observations have proposed the existence of distinctive biochemical characteristics of the amygdala-predominant alpha-synuclein aggregates, which may be more informative regarding the pathogenic interaction of tau and alpha-synuclein (Ferman et al., 2020; Nelson et al., 2018; Sorrentino et al., 2019).

Our study showed that except for CAA and LBD, the most common copathologies of AD, namely TDP-43 proteinopathy, AGD, HS and VBI are, as expected, more common and severe in LOAD as opposed to EOAD (Karanth et al., 2020; Robinson et al., 2018; Rodriguez et al., 2016). A higher prevalence of TDP-43 pathology and HS has been described in LOAD compared to EOAD (Davidson et al., 2011). ARTAG was more commonly seen in LOAD cases compared to EOAD, as expected (Kovacs et al., 2016). ATAC pathology has been associated with worse performance on neuropsychological scores in domains corresponding to the function of the anatomical area affected by this type of pathology (Resende et al., 2020). These findings may have implications regarding the atypical, non-amnestic clinical presentations of AD, which are more common in EOAD than in LOAD (Resende et al., 2020). While in our study ATAC were more commonly found in EOAD than in LOAD, the difference did not reach statistical significance, perhaps in view of the high prevalence of non-amnestic AD presentations in our LOAD cohort. Our study found a significant association between ApoE e4 and a higher number of copathologies, but no significant association between ApoE e4 and any of the specific pathologies, except for a trend for AGD. This is likely a consequence of the cohort size, also possibly secondary to the high prevalence in both cohorts of patients with non-amnestic AD clinical presentations (Balasa et al., 2011; van der Flier et al., 2011).

A more rapid clinic decline has been associated with a higher severity of neuritic plaque and neurofibrillary tangles pathology in EOAD compared to LOAD (Ho et al., 2002; Middleton et al., 2011). This is consistent with recent observations based on the use of in vivo tau-PET demonstrating higher tau-PET signal in EOAD compared to LOAD, a difference that is significantly associated with faster rate of atrophy in EOAD compared to LOAD (La Joie et al., 2020). In our study we observed higher severity of AD neuropathological hallmarks in EOAD compared to LOAD and higher rate of disease progression. There was a significantly longer survival from disease onset in EOAD compared to LOAD, a finding previously described, and possibly linked to the overall better systemic health status in the younger cohort (Gronning et al., 2012). Longer survival may, in part allow for larger pathology burden to accumulate over time. On the other hand, differences in rate of proteinopathy accumulation have been described in various AD subtypes, and are likely to play a major role in the expression of the clinical phenotype (Murray et al., 2011).

Our study presents some limitations. While we report findings on a large autopsy series of EOAD cases, the overall cohort of AD cases remains relatively small, therefore increasing the chances of occurrence of selection biases at the time of enrollment. Our autopsy cohort consists of research participants referred from tertiary care, academic centers and maybe not reflecting of the prevalence of brain copathologies in a community-dwelling population. The proportion of cases with atypical, non-amnestic, AD presentation is higher in our cohort than in the general population. Therefore our findings may not be fully representative of the differences between EOAD and LOAD patients with amnestic AD. Finally, our protocol for assessing VBI is likely to underestimate the prevalence of this co-pathology.

In summary, our study shows that non-AD neuropathology is common in sporadic EOAD despite a lower prevalence than in LOAD. The number of copathologies predicted worse cognitive performance in both cohorts. Coexistent LBD pathology was as frequent in EOAD as in LOAD, particularly in the amygdala-predominant variant. Our findings suggest that the role of copathology should be considered when assessing EOAD clinical phenotype and response to treatment both in clinical practice and in clinical trials. In an era when in vivo PET (Fleisher et al., 2020; La Joie et al., 2019; Lowe et al., 2019) (and Soleimani et al. Brain 2020, In press) and plasma biomarkers (Janelidze et al., 2020; Rodriguez et al., 2020; Thijssen et al., 2020) can accurately identify cases with intermediate to high ADNC with high sensitivity and specificity, it will be critical to remain mindful that additional copathologies, for which no robust biomarker exists today, are likely to be present even in patients with younger disease onset.

## Data Availability

All data used in this study are available for review upon request.

## Acknowledgments

We are indebted to the research participants and their families for their generous contribution to science.

## Supplementary Materials

**eFigure 1.**
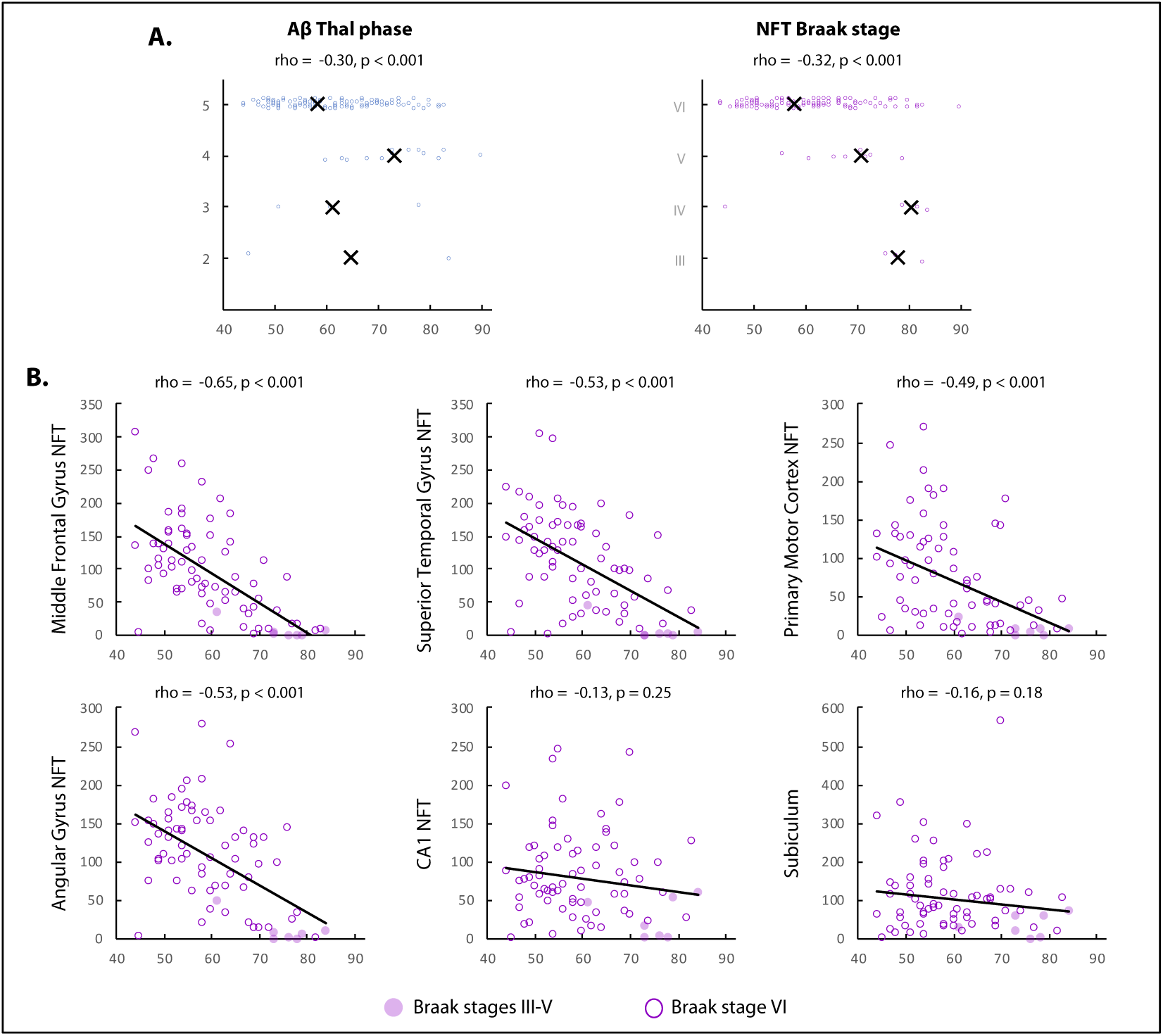
Equivalent to Figure 2, with age on onset treated as a continuous variable. Analyses are equivalent to Figure 2 but considering age at symptom onset as a continuous variable instead of splitting cases based on a 65yo threshold. In panel A, black crosses indicate the median ages for each level of pathology. In panel B, cases with Braak stage VI are highlighted to emphasize the variability of NFT burden despite ceiling Braak staging.

**eFigure 2.**
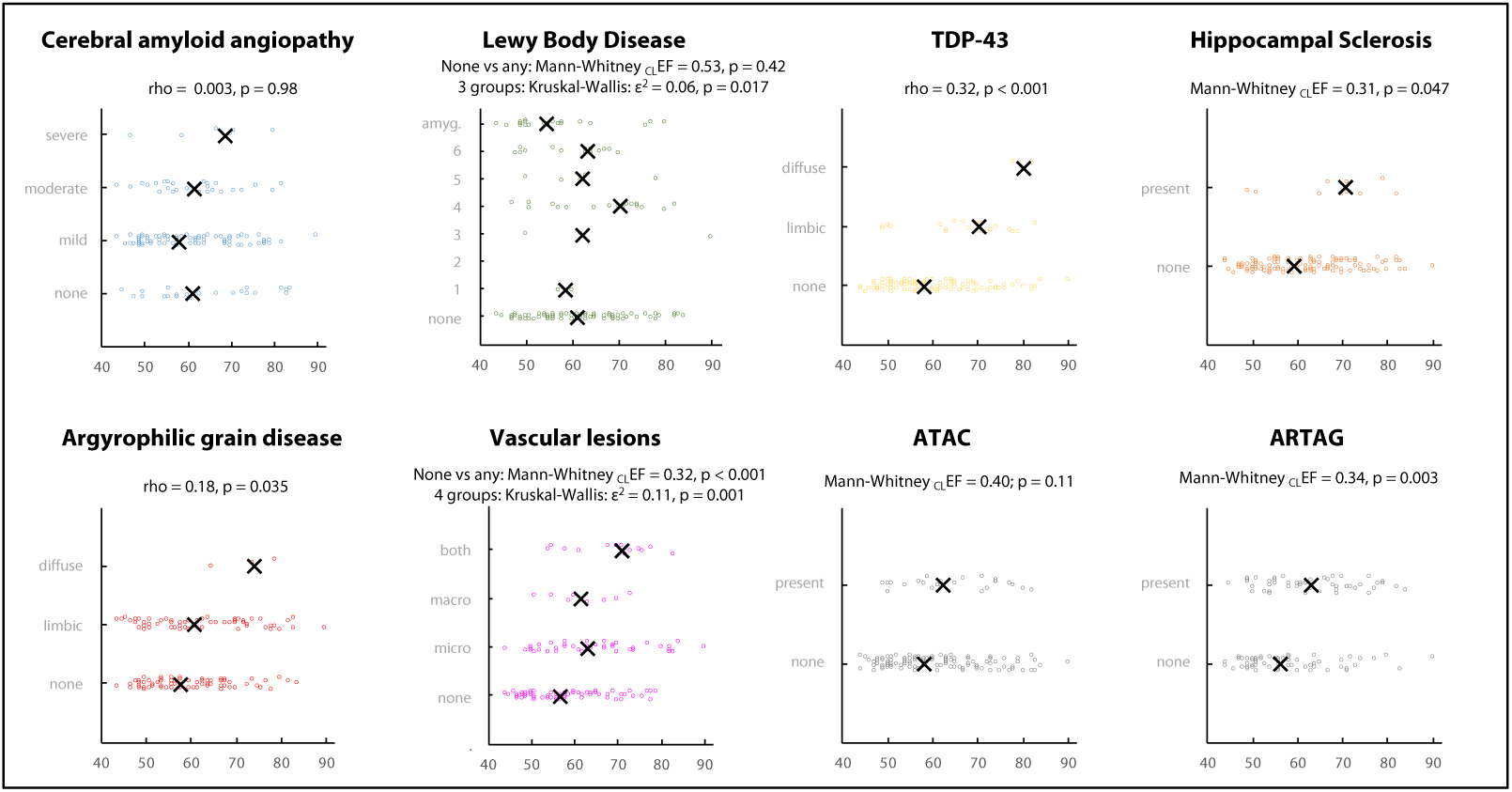
Age of symptom onset and specific coexistent non-AD pathologies. Analyses are equivalent to Figure 4 but considering age at symptom onset as a continuous variable instead of splitting cases based on a 65yo threshold. Black crosses indicate the median ages for each level of pathology.

**eFigure 3.**
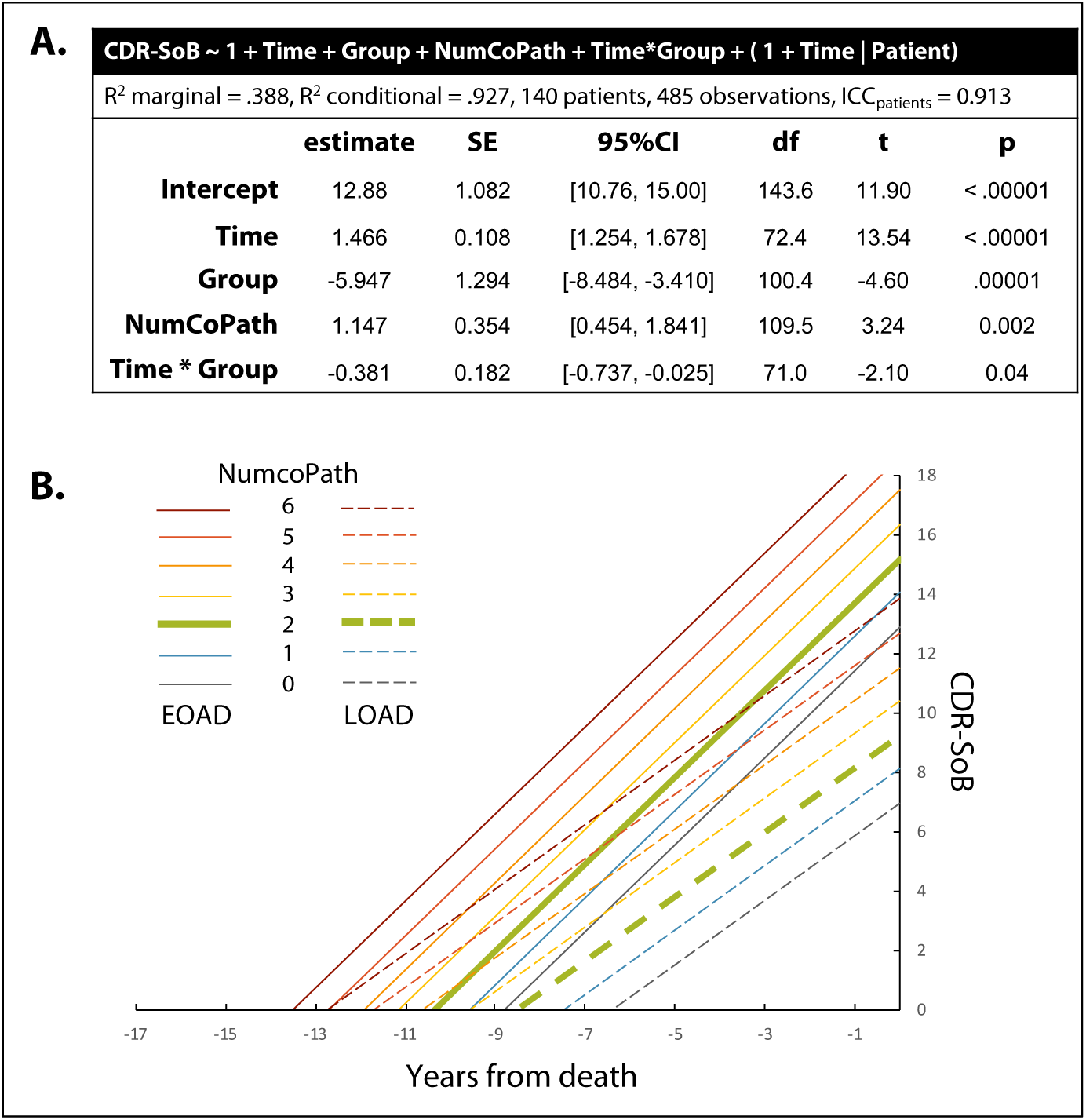
Pre mortem cognitive decline based on CDR-SoB. A. Optimal linear mixed effect model as identified in eTable4. B. CDR-SoB trajectories as predicted by model in B. The trajectories for EOAD and LOAD patients with 2 copathologies (the median number of copathologies in the whole cohort) are emphasized.

**eTable 1.** Multivariable regression analyses assessing the independent contribution of age of onset group, sex, and disease duration on Neurofibrillary Tangle density.

|  | Std estimate | p | R<sup>2</sup> model |
| --- | --- | --- | --- |
| **Middle Frontal Gyrus NFT** |  |  | 0.43 |
| LOAD vs EOAD | -1.27 | <0.001 |  |
| Male vs Female | -0.47 | 0.01 |  |
| Disease Duration | -0.04 | 0.68 |  |
| **Superior Temporal Gyrus NFT** |  |  | 0.39 |
| LOAD vs EOAD | -0.91 | <0.001 |  |
| Male vs Female | -0.81 | <0.001 |  |
| Disease Duration | 0.03 | 0.75 |  |
| **Primary Motor Cortex NFT** |  |  | 0.20 |
| LOAD vs EOAD | -0.77 | 0.002 |  |
| Male vs Female | -0.44 | 0.047 |  |
| Disease Duration | -0.06 | 0.57 |  |
| **Angular Gyrus NFT** |  |  | 0.37 |
| LOAD vs EOAD | -1.06 | <0.001 |  |
| Male vs Female | -0.62 | 0.002 |  |
| Disease Duration | -0.02 | 0.82 |  |
| **CA1 NFT** |  |  | 0.21 |
| LOAD vs EOAD | 0.05 | 0.82 |  |
| Male vs Female | -0.51 | 0.02 |  |
| Disease Duration | 0.38 | <0.001 |  |
| **Subiculum NFT** |  |  | 0.16 |
| LOAD vs EOAD | 0.06 | 0.81 |  |
| Male vs Female | -0.55 | 0.01 |  |
| Disease Duration | 0.30 | 0.01 |  |

Positive estimates correspond to higher NFT density in LOAD (compared to EOAD), in Males (compared to Females), or in patients with a longer disease duration.

**eTable 2.** Associations of APOE4 and age of onset on Neurofibrillary Tangle density.

|  | estimate | 95%CI | Std estimate | p |
| --- | --- | --- | --- | --- |
| **Middle Frontal Gyrus NFT (n=71)** |  |  |  |  |
| APOE4 (33 vs 38) | 7.19 | [-20.39, 34.77] | 0.099 | 0.61 |
| Age of onset | -4.52 | [-5.93, -3.12] | -0.614 | < .001 |
| **Superior Temporal Gyrus NFT (n=70)** |  |  |  |  |
| APOE4 (33 vs 37) | -1.10 | [-30.19, 27.99] | -0.016 | 0.94 |
| Age of onset | -3.89 | [-5.42, -2.36] | -0.527 | < .001 |
| **Primary Motor Cortex NFT (n=67)** |  |  |  |  |
| APOE4 (32 vs 35) | -23.90 | [-52.82, 5.03] | -0.358 | 0.10 |
| Age of onset | -3.01 | [-4.46, -1.56] | -0.452 | < .001 |
| **Angular Gyrus NFT (n=70)** |  |  |  |  |
| APOE4 (33 vs 37) | -0.44 | [-27.9, 27.02] | -0.007 | 0.97 |
| Age of onset | -3.51 | [-4.91, -2.10] | -0.521 | < .001 |
| **CA1 NFT (n=70)** |  |  |  |  |
| APOE4 (33 vs 37) | 10.89 | [-16.20, 37.97] | 0.193 | 0.43 |
| Age of onset | -0.63 | [-2.04, 0.77] | -0.109 | 0.37 |
| **Subiculum NFT (n=69)** |  |  |  |  |
| APOE4 (33 vs 36) | 30.99 | [-16.36, 78.34] | 0.314 | 0.20 |
| Age of onset | -1.02 | [-3.48, 1.44] | -0.100 | 0.41 |

APOE4 is coded as e4 carrier versus non-carrier, the latter being the model reference (numbers in brackets indicate e4 carriers versus non-carriers). Age of onset is a continuous variable, expressed in years.

**eTable 3.** Linear Mixed effect Models (dependent variable: MMSE): model selection.

| model | 1 | 2 | 3 | 4 | 5 | 6 | 7 | 8 | 9 | 10 | 11 | 12 | 13 | 14 |
| --- | --- | --- | --- | --- | --- | --- | --- | --- | --- | --- | --- | --- | --- | --- |
| Time till death (-17.7 to -0.2 y) | X | X | X | X | X | X | **X** | X | X | X | X | X | X | X |
| Group (EOAD/LOAD) |  | X |  | X | X |  | **X** | X | X | X | X | X | X | X |
| NumCoPath (0 to 6) |  |  | X | X |  | X | **X** | X | X | X | X | X | X | X |
| Time * Group |  |  |  |  | X |  | **X** |  |  | X | X |  | X | X |
| Time * NumCoPath |  |  |  |  |  | X |  | X |  | X |  | X | X | X |
| Group * NumCoPath |  |  |  |  |  |  |  |  | X |  | X | X | X | X |
| Time * Group * NumCopath |  |  |  |  |  |  |  |  |  |  |  |  |  | X |
| AIC | 2391.1 | 2378.5 | 2392.4 | 2375.7 | 2365.8 | 2393.8 | **2362.9** | 2377.5 | 2376.7 | 2364.7 | 2362.7 | 2378.4 | 2364.5 | 2365.4 |
| ΔAIC | 28.4 | 15.8 | 29.7 | 13 | 3.1 | 31.1 | 0.2 | 14.8 | 14 | 2 | 0 | 15.7 | 1.8 | 2.7 |
| BIC | 2415.1 | 2403.9 | 2419.5 | 2404.3 | 2395.4 | 2426.8 | **2395.6** | 2412 | 2407 | 2403.5 | 2397.1 | 2414.7 | 2405 | 2410.6 |
| ΔBIC | 19.7 | 8.5 | 24.1 | 8.9 | 0 | 31.4 | 0.2 | 16.6 | 11.6 | 8.1 | 1.7 | 19.3 | 9.6 | 15.2 |

NumCoPath: Number of copathologies, AIC: Akaike Information Criterion, BIC: Bayesian Information Criterion, $\Delta$ AIC/BIC: difference between each model and the optimal model (with lowest AIC/BIC).
All models included a fixed effect for time until death (in years) and random intercept and slope. AIC and BIC disagreed on which model was optimal, with AIC pointing to model 11 while BIC pointed to a more parsimonious model (#4). Overall, Model 7 was chosen as a compromise given that it was the converging third best model according to both indices and showed acceptable change compared to the best models ( $\Delta$ AIC = 0.2, $\Delta$ BIC = 0.2).

**eTable 4.** Linear Mixed effect Models: model selection (dependent variable: CDR-SoB)

| model | 1 | 2 | 3 | 4 | 5 | 6 | 7 | 8 | 9 | 10 | 11 | 12 | 13 | 14 |
| --- | --- | --- | --- | --- | --- | --- | --- | --- | --- | --- | --- | --- | --- | --- |
| Time till death (-17.7 to -0.06 y) | X | X | X | X | X | X | **X** | X | X | X | X | X | X | X |
| Group (EOAD/LOAD) |  | X |  | X | X |  | **X** | X | X | X | X | X | X | X |
| NumCoPath (0 to 6) |  |  | X | X |  | X | **X** | X | X | X | X | X | X | X |
| Time * Group |  |  |  |  | X |  | **X** |  |  | X | X |  | X | X |
| Time * NumCoPath |  |  |  |  |  | X |  | X |  | X |  | X | X | X |
| Group * NumCoPath |  |  |  |  |  |  |  |  | X |  | X | X | X | X |
| Time * Group * NumCoPath |  |  |  |  |  |  |  |  |  |  |  |  |  | X |
| AIC | 2518.3 | 2509.6 | 2516.6 | 2501.2 | 2507 | 2517.1 | **2498.9** | 2501.7 | 2501 | 2497.9 | 2497.9 | 2501.4 | 2496.6 | 2494.3 |
| ΔAIC | 24 | 15.3 | 22.3 | 6.9 | 12.7 | 22.8 | 4.6 | 7.4 | 6.7 | 3.6 | 3.6 | 7.1 | 2.3 | 0 |
| BIC | 2546.2 | 2540.1 | 2549 | 2536.3 | 2543.3 | 2557.2 | **2539.8** | 2544.6 | 2539.1 | 2546.6 | 2541.9 | 2547.3 | 2548.4 | 2552.9 |
| ΔBIC | 9.9 | 3.8 | 12.7 | 0 | 7 | 20.9 | 3.5 | 8.3 | 2.8 | 10.3 | 5.6 | 11 | 12.1 | 16.6 |

NumCoPath: Number of copathologies, AIC: Akaike Information Criterion, BIC: Bayesian Information Criterion, $\Delta$ AIC/BIC: difference between each model and the optimal model (with lowest AIC/BIC).
All models included a fixed effect for time until death (in years) and random intercept and slope. AIC and BIC disagreed on which model was optimal, with AIC pointing to the most complex model (#14, including all simple effects and interactions), while BIC pointed to a more parsimonious model (#4, with all three simple effects but no interaction). Overall, Model 7 was chosen as a compromise given that it showed acceptable change compared to the best models ( $\Delta$ AIC = 4.6, $\Delta$ BIC = 3.5).

---

STROBE Statement—checklist of items that should be included in reports of observational studies

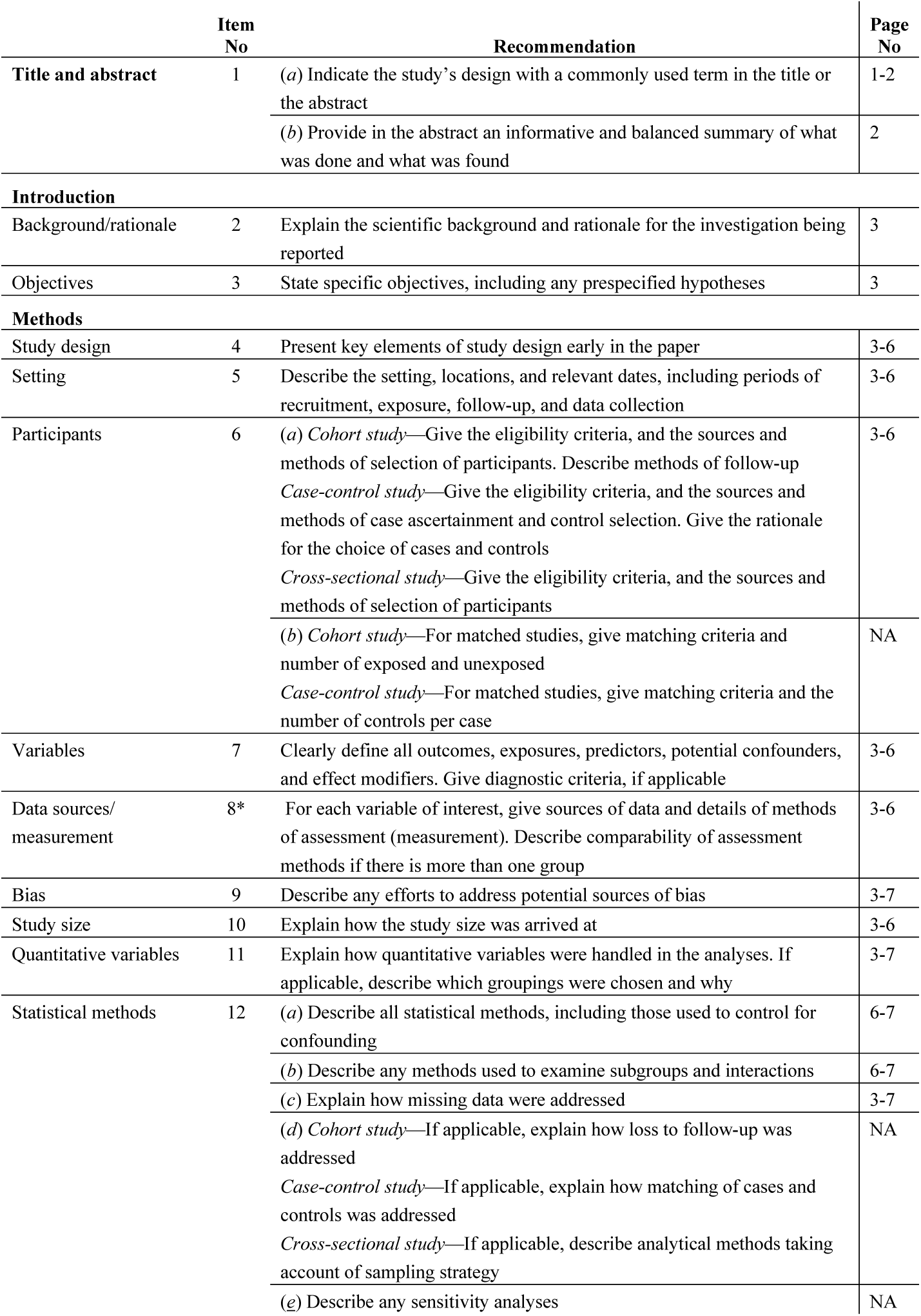
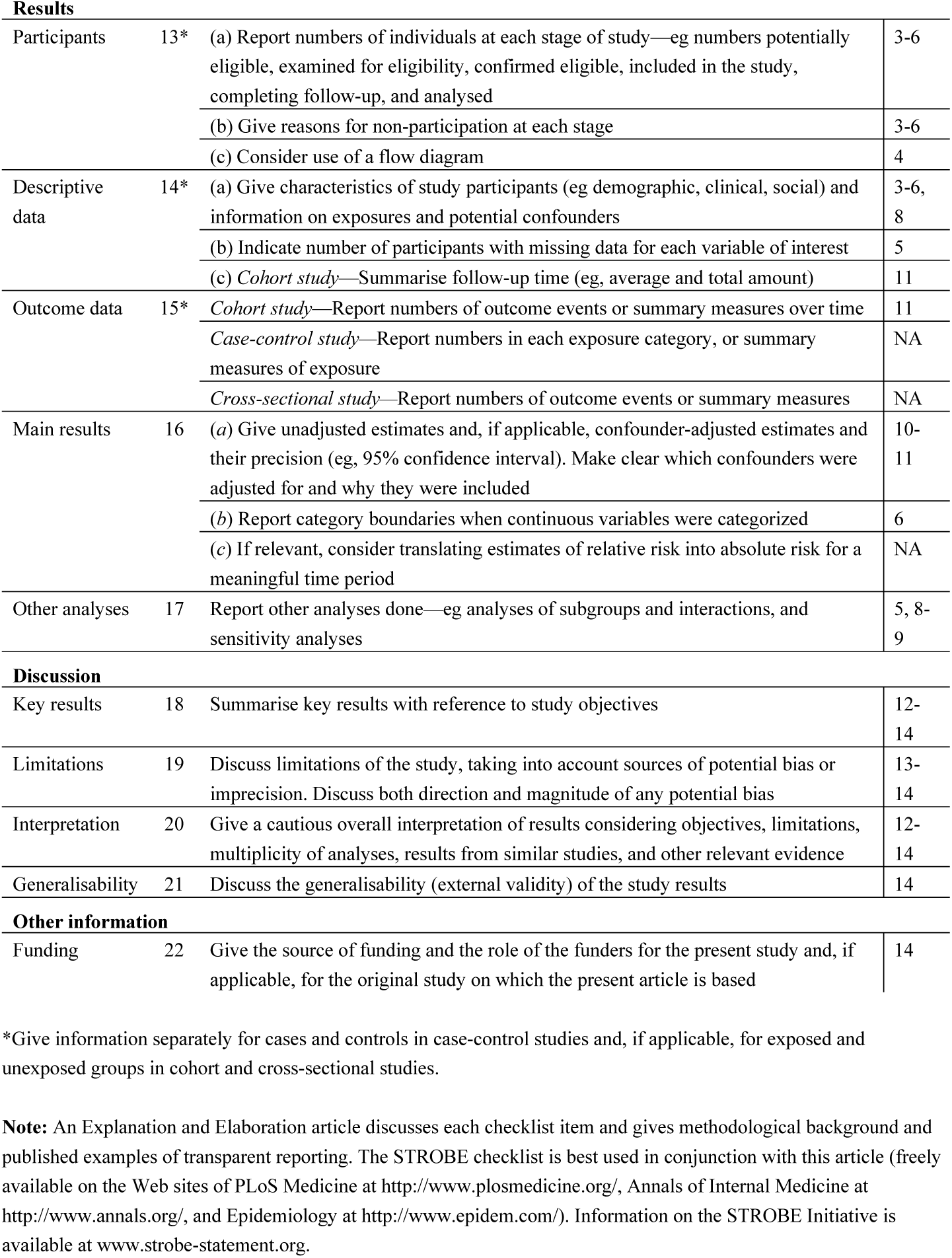

|  | Item No | Recommendation | Page No |
| --- | --- | --- | --- |
| Title and abstract | 1 | (a) Indicate the study’s design with a commonly used term in the title or the abstract | 1-2 |
|  |  | (b) Provide in the abstract an informative and balanced summary of what was done and what was found | 2 |
| Introduction |  |  |  |
| Background/rationale | 2 | Explain the scientific background and rationale for the investigation being reported | 3 |
| Objectives | 3 | State specific objectives, including any prespecified hypotheses | 3 |
| Methods |  |  |  |
| Study design | 4 | Present key elements of study design early in the paper | 3-6 |
| Setting | 5 | Describe the setting, locations, and relevant dates, including periods of recruitment, exposure, follow-up, and data collection | 3-6 |
| Participants | 6 | (a) Cohort study—Give the eligibility criteria, and the sources and methods of selection of participants. Describe methods of follow-up<br>Case-control study—Give the eligibility criteria, and the sources and methods of case ascertainment and control selection. Give the rationale for the choice of cases and controls<br>Cross-sectional study—Give the eligibility criteria, and the sources and methods of selection of participants | 3-6 |
|  |  | (b) Cohort study—For matched studies, give matching criteria and number of exposed and unexposed<br>Case-control study—For matched studies, give matching criteria and the number of controls per case | NA |
| Variables | 7 | Clearly define all outcomes, exposures, predictors, potential confounders, and effect modifiers. Give diagnostic criteria, if applicable | 3-6 |
| Data sources/<br>measurement | 8* | For each variable of interest, give sources of data and details of methods of assessment (measurement). Describe comparability of assessment methods if there is more than one group | 3-6 |
| Bias | 9 | Describe any efforts to address potential sources of bias | 3-7 |
| Study size | 10 | Explain how the study size was arrived at | 3-6 |
| Quantitative variables | 11 | Explain how quantitative variables were handled in the analyses. If applicable, describe which groupings were chosen and why | 3-7 |
| Statistical methods | 12 | (a) Describe all statistical methods, including those used to control for confounding | 6-7 |
|  |  | (b) Describe any methods used to examine subgroups and interactions | 6-7 |
|  |  | (c) Explain how missing data were addressed | 3-7 |
|  |  | (d) Cohort study—If applicable, explain how loss to follow-up was addressed<br>Case-control study—If applicable, explain how matching of cases and controls was addressed<br>Cross-sectional study—If applicable, describe analytical methods taking account of sampling strategy | NA |
|  |  | (e) Describe any sensitivity analyses | NA |
| **Results** |  |  |  |
| Participants | 13* | (a) Report numbers of individuals at each stage of study—eg numbers potentially eligible, examined for eligibility, confirmed eligible, included in the study, completing follow-up, and analysed | 3-6 |
|  |  | (b) Give reasons for non-participation at each stage | 3-6 |
|  |  | (c) Consider use of a flow diagram | 4 |
| Descriptive data | 14* | (a) Give characteristics of study participants (eg demographic, clinical, social) and information on exposures and potential confounders | 3-6, 8 |
|  |  | (b) Indicate number of participants with missing data for each variable of interest | 5 |
|  |  | (c) *Cohort study*—Summarise follow-up time (eg, average and total amount) | 11 |
| Outcome data | 15* | *Cohort study*—Report numbers of outcome events or summary measures over time | 11 |
|  |  | *Case-control study*—Report numbers in each exposure category, or summary measures of exposure | NA |
|  |  | *Cross-sectional study*—Report numbers of outcome events or summary measures | NA |
| Main results | 16 | (a) Give unadjusted estimates and, if applicable, confounder-adjusted estimates and their precision (eg, 95% confidence interval). Make clear which confounders were adjusted for and why they were included | 10-11 |
|  |  | (b) Report category boundaries when continuous variables were categorized | 6 |
|  |  | (c) If relevant, consider translating estimates of relative risk into absolute risk for a meaningful time period | NA |
| Other analyses | 17 | Report other analyses done—eg analyses of subgroups and interactions, and sensitivity analyses | 5, 8-9 |
| **Discussion** |  |  |  |
| Key results | 18 | Summarise key results with reference to study objectives | 12-14 |
| Limitations | 19 | Discuss limitations of the study, taking into account sources of potential bias or imprecision. Discuss both direction and magnitude of any potential bias | 13-14 |
| Interpretation | 20 | Give a cautious overall interpretation of results considering objectives, limitations, multiplicity of analyses, results from similar studies, and other relevant evidence | 12-14 |
| Generalisability | 21 | Discuss the generalisability (external validity) of the study results | 14 |
| **Other information** |  |  |  |
| Funding | 22 | Give the source of funding and the role of the funders for the present study and, if applicable, for the original study on which the present article is based | 14 |

\*Give information separately for cases and controls in case-control studies and, if applicable, for exposed and unexposed groups in cohort and cross-sectional studies.
**Note:** An Explanation and Elaboration article discusses each checklist item and gives methodological background and published examples of transparent reporting. The STROBE checklist is best used in conjunction with this article (freely available on the Web sites of PLoS Medicine at http://www.plosmedicine.org/, Annals of Internal Medicine at http://www.annals.org/, and Epidemiology at http://www.epidem.com/). Information on the STROBE Initiative is available at www.strobe-statement.org.

